## Supplemental file for "Doxycycline for the prevention of progression of COVID-19 to severe disease requiring intensive care unit (ICU) admission: a randomized, controlled, open-label, parallel group trial (DOXPREVENT.ICU)"

**Supplemental Appendix**

**1. Sample size calculation**

(A) Original sample size calculation (per protocol)

### Sample size justification

We assumed 25% of patients newly admitted to hospital with COVID-19 will require ICU care within 14 days of admission, and that the addition of 200mg doxycycline qd to standard of care will reduce the need for ICU transfer by 50%, a risk ratio of 0.500. Loss to follow up was assumed to be no more than 5%. A 2:1 randomisation ratio (Doxycycline plus SoC : SoC) was to be used.

We required 80% power using a one-sided alpha of 2.5%.

The sample size required for a conventional trial, with no interim analyses, looking at the difference in the rate of transfer to ICU was 220 in the doxycycline group and 110 in the SoC group, a total of 330. Allowing for a 5% dropout rate, the corresponding figures are 231, 116 and 347.

### Interim analyses and stopping rules

Interim analyses allowing the possibility of stopping for both success and futility was to occur once the status of 50% and 75% of participants was known. In addition, a futility-only interim was to occur once the status of 25% of participants was known. A binding O’Brien-Fleming-like alpha spending function will be used to define the success boundary and a Pockock-like beta spending function will define the futility boundary.

The design characteristics of this design are summarised in the table below.

| ***Operating characteristics of the chosen design*** | | | | |
| --- | --- | --- | --- | --- |
|  | ***Interim 1*** | ***Interim 2*** | ***Interim 3*** | ***Final*** |
| Information rate | 25.0% | 50.0% | 75.0% | 100.0% |
| Total sample size* | 110 | 220 | 329 | 439 |
| Cumulative alpha spent | 0.0000 | 0.0015 | 0.0092 | 0.0220 |
| Cumulative power | 0.000 | 0.250 | 0.649 | 0.800 |
| One-sided local significance level | 0 | 0.00153 | 0.00866 | 0.0189 |
| Efficacy boundary (Z-value scale) | NA | 2.963 | 2.380 | 2.078 |
| Efficacy boundary (approximate treatment effect scale) | NA | -0.151 | -0.107 | -0.083 |
| Futility boundary (Z-value scale) | 0.153 | 0.933 | 1.531 | NA |
| Futility boundary (approximate treatment effect scale) | -0.013 | -0.055 | -0.072 | NA |
| Overall exit probability (under H1) | 0.071 | 0.302 | 0.441 | NA |
| Exit probability for efficacy (under H1) | 0.000 | 0.250 | 0.399 | 0.151 |
| Exit probability for futility (under H1) | 0.071 | 0.053 | 0.042 | NA |
| Overall exit probability (under H0) | 0.561 | 0.287 | 0.112 | NA |
| Exit probability for efficacy (under H0) | 0.000 | 0.002 | 0.007 | 0.008 |
| Exit probability for futility (under H0) | 0.561 | 0.286 | 0.104 | NA |
| *: Evaluable patients, without allowance for drop outs | | | | |

Specifically, the (approximate) values of observed treatment effect that would trigger a recommendation to stop the study at each analysis are given in the table below.

| ***Approximate boundary values on the treatment effect scale*** | | |
| --- | --- | --- |
| ***Analysis*** | ***Futility*** | ***Success*** |
| Interim 1 | -1.3% |  |
| Interim 2 | -5.5% | -15.1% |
| Interim 3 | -7.2% | -10.7% |
| Final |  | -8.3% |

For example, at the second interim, if the observed treatment effect is greater than -5.5%, the study will stop for futility. If it is less than -15.1%, it will stop for success. Otherwise, it will continue.

The maximum number of participants required using this design is 439, an increase of 109, before allowing for dropouts, compared to the conventional design. However, the expected sample size is reduced to 300.4 when doxycycline has the desired effect on ICU transfer and 178.9 when it has no effect.

It was thought unlikely that interims will occur precisely at the information fractions given above. Consequently, boundary values for subsequent interims were to be adjusted to ensure that the operating characteristics of the design are preserved. The adjusted boundary values for future interims would be documented in writing in the report that summarises the results of each interim analysis.

Some other properties of the design are summarised in the graphs below.

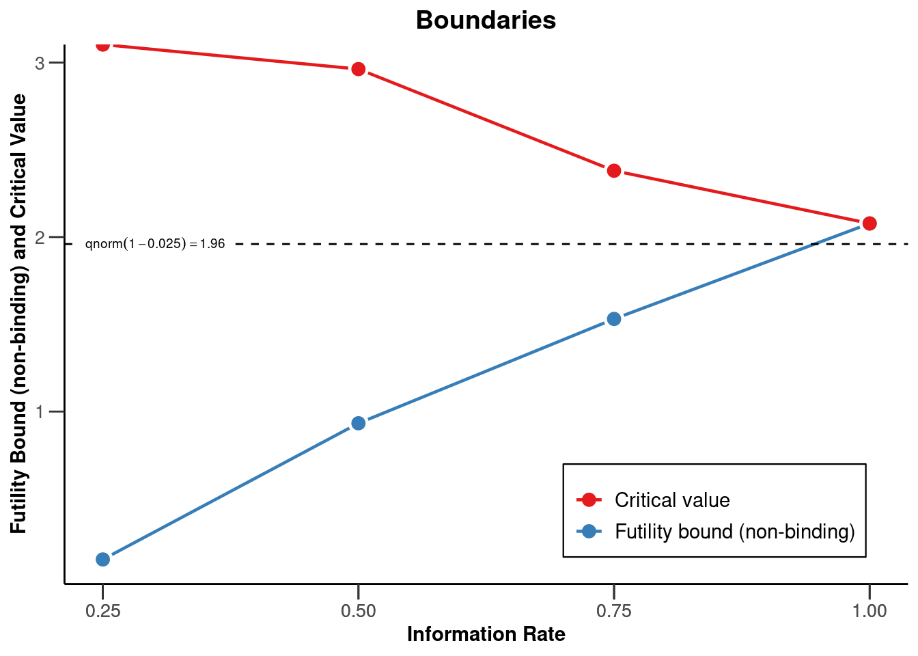

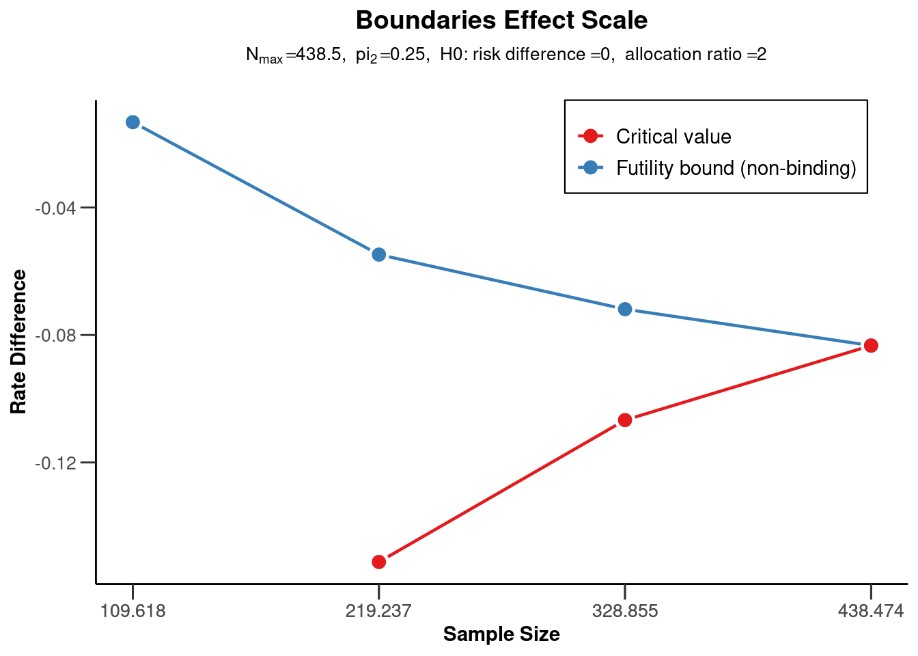

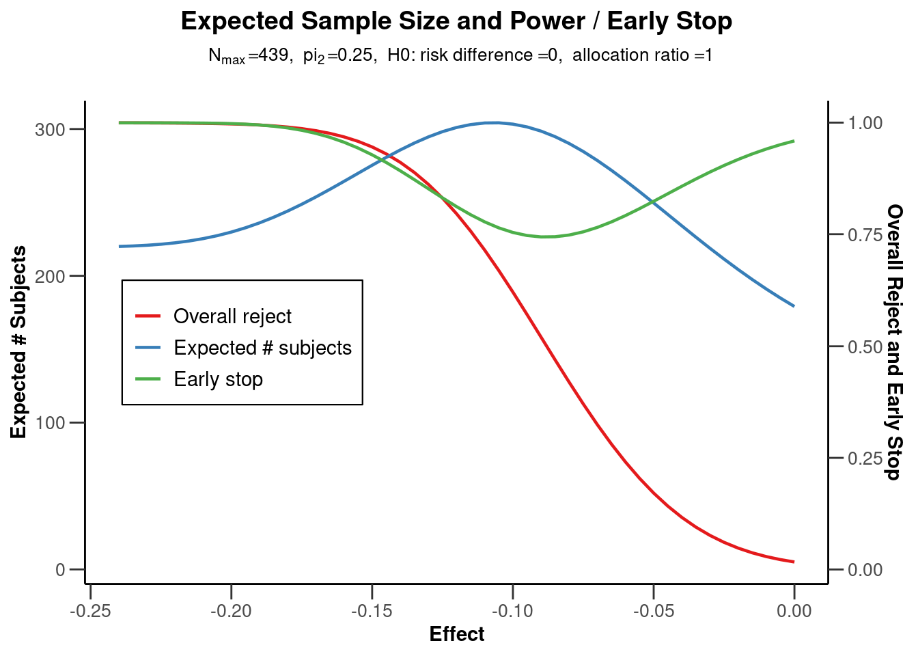

(B) Actual sample size calculation

Due to an error in programming, the study app was programmed for 1:1 randomisation.

We assumed that 25% of patients newly admitted to hospital with COVID-19 will require ICU care within 14 days of admission. The addition of 100mg doxycycline BID to standard of care will reduce the need for ICU transfer within by 50%, a risk ratio of 0.500. Loss to follow up will be no more than 5%. A 1:1 randomisation ratio (SoC+Doxy: SoC) will be used. Loss to follow up was assumed to be ≤5%.

We required 80% power using a one-sided alpha of 2.5%. The sample size required for a conventional trial, with no interim analyses, looking at the difference in the rate of transfer to ICU was 152 in the SoC+doxy arm and 152 in the SoC arm, a total of 304. Allowing for a 5% dropout rate, the corresponding figures were 160, 160 and 320. As we had already exceeded this number of evaluable subjects, no additional calculations to implement a group sequential approach were required.

**Supplementary Table 1. Number of Risk factors (RF; comorbidities associated with risk for developing severe COVID-19) in patients randomized to SoC and SoC+Doxy groups.**

| **RF (n)** | **Randomised to SoC+Doxy (n=192)** | | **Randomised to SoC (n=195)** | |
| --- | --- | --- | --- | --- |
|  | **n with RF** | **RF total (n)** | **n with RF** | **RF total (n)** |
| 0 | 48 | 0 | 49 | 0 |
| 2 | 3 | 6 | 7 | 14 |
| 3 | 44 | 132 | 44 | 132 |
| 4 | 55 | 220 | 52 | 208 |
| 5 | 40 | 200 | 34 | 170 |
| 6 | 2 | 12 | 8 | 48 |
| 7 | 0 | 0 | 1 | 7 |
|  | | **Total RF (n) = 570** |  | **Total RF (n) = 579** |
|  |  | average/all = 3.0 |  | average/all = 3.0 |
|  |  | average/RF+ = 4.0 |  | average/RF+ = 4.0 |

**Supplementary Figure 1.**

**Effect of doxycycline on SARS-CoV-2 infection of ACE-2-overexpressing lung-derived cell lines: Doxycycline and Tetracycline have no effect on SARS-CoV-2 replication.**

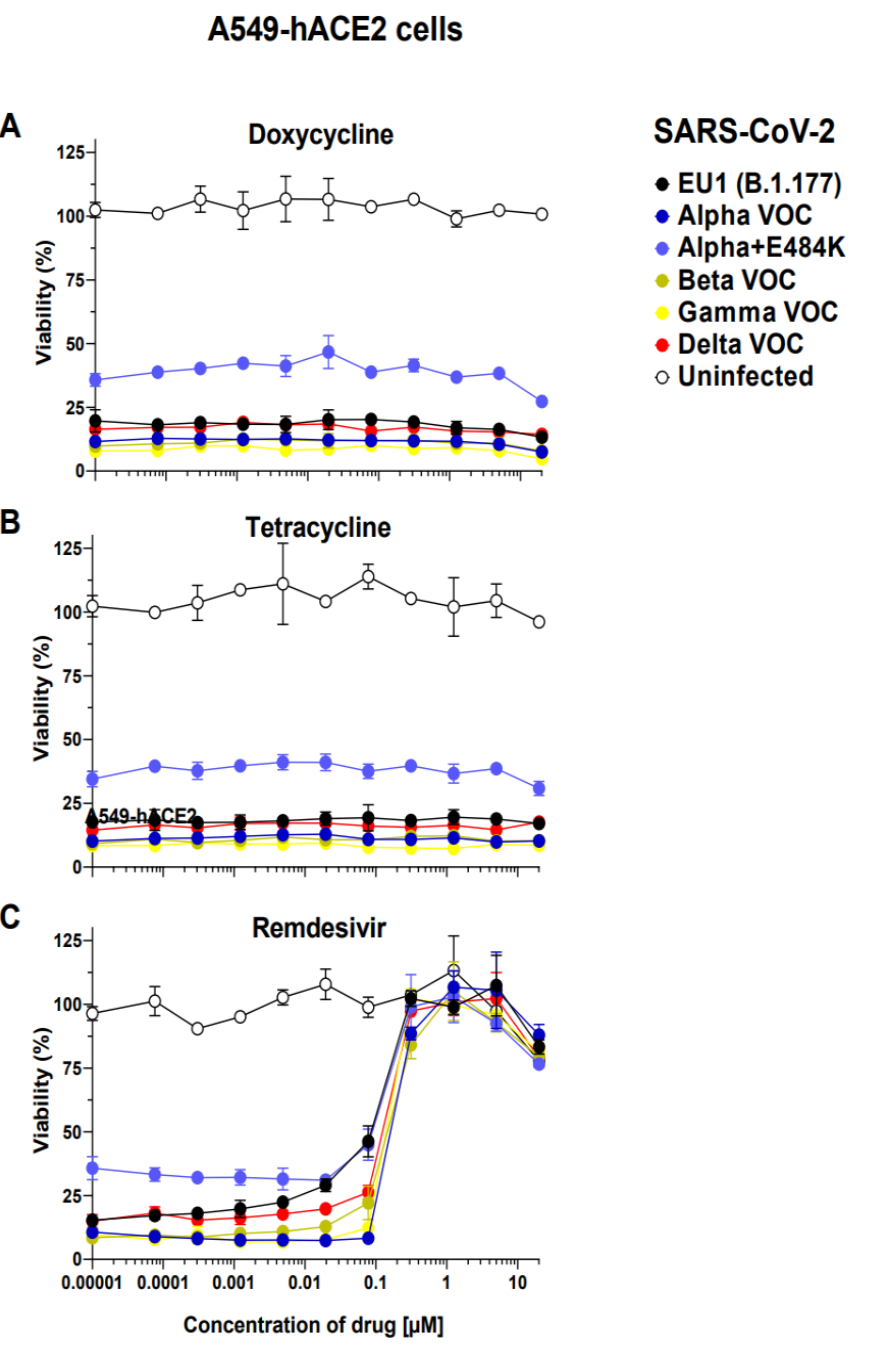

**Supplementary Figure 1. Legend:** Human adenocarcinomic alveolar basal epithelial A549-hACE2 cells were treated with serial dilutions of the indicated drugs (doxycycline, tetracycline, remdesivir) starting six hours before challenge with the indicated SARS-CoV-2 variants. The highest drug concentration on cells was 16.7 µM and drug treatment was continued until cell harvest. 72 hrs after infection, viability of target cells was determined. Error bars represent mean and standard deviation of four (virus) or two (uninfected) technical replicates. The EC50 values for remdesivir ranged from 70 to 151 nM for the different SARS-CoV-2 variants.

**Supplementary Figure 1. Material and Methods.**

Cell culture. Vero-E6 cells (American Type Culture Collection, ATCC, Virginia, USA), A549 cells (human adenocarcinomic alveolar basal epithelial cells; American Type Culture Collection, ATCC, Virginia, USA), Caco-2 cells (human colorectal adenocarcinoma cell line, ATCC, Virginia, USA) and MDA-MB-231 cells (triple-negative human breast adenocarcinoma; DSMZ-German Collection of Microorganisms and Cell Cultures GmbH, Braunschweig, Germany) were cultivated at 37°C in a humidified incubator with atmospheric oxygen concentrations (21 %) and 5 % CO2. Cells were maintained in Dulbecco’s Modified Eagle’s Medium (DMEM) high glucose containing 10% fetal bovine serum (FBS), 100 U/mL penicillin-streptomycin and NEAA (culture medium). Cells were routinely passaged when reaching a confluence of 80 - 90%.

Isolation and expansion of SARS-CoV-2 clinical isolates. Caco-2 cells cultivated in “virus isolation medium” (DMEM, 2% FBS, 100 U/mL penicillin-streptomycin, NEAA, 0.5 µg/mL gentamicin, and 0.25 µg/mL amphotericin B) were challenged for 2 h with a clinical isolate of the B.1.177 (EU1) lineage (GISAID EPI ISL: 3233461) obtained from a nasopharyngeal swab of a COVID-19 patient. Subsequently, the virus isolation medium was replaced with regular culture medium, and three days post infection supernatant was collected and passaged onto Vero-E6 cells (ATCC) in virus isolation medium. After three additional days, cell culture supernatants were harvested and stored at -80°C. Further expansion of viruses was performed in “virus expansion medium” (DMEM containing 5% FBS, 100 U/mL penicillin-streptomycin, NEAA). VOCs Alpha (B.1.1.7; GISAID EPI ISL: 3233462), Beta (B.1.351; (GISAID EPI ISL: 1752394), Gamma (P.1 / B.1.1.28.1; (GISAID EPI ISL: 2095178)) and Delta (B.1.617.2; (GISAID EPI ISL: 2772700)) were kindly provided by the Bavarian Landesamt für Gesundheit und Lebensmittelsicherheit (LGL). VOC Alpha + E484K (B.1.1.7 + E484K; GISAID EPI ISL: 2772697) was obtained from the Institute for Virology, Innsbruck, Austria. All VOCs were expanded in expansion medium on Vero-E6 cells. Virus stocks were characterized by RT-qPCR, as reported previously (1). In parallel, for expanded stocks of SARS-CoV-2 near full-length genome sequences were obtained following the ARTIC network nCoV-2019 sequencing protocol v2 (2) as described previously (3).

Infection of A549-hACE2 cells and MDA-MB-231-hACE2 cells with SARS-CoV-2 – drug screening viability assay. A549-hACE2 cells (7.5 x 103 cells per well) and MDA-MB-231-hACE2 cells (1.0 x 104 cells per well) were plated in a 384-well white well plate (Corning) in “virus infection medium” (DMEM, 2% FBS, 100 U/mL penicillin-streptomycin, NEAA). Target cells were treated with either medium or a serial dilution of remdesivir (Adooq Biosciences), doxycycline (Sigma-Aldrich) or tetracycline (Sigma-Aldrich) six hours before infection. Subsequently, cells were challenged with the indicated SARS-CoV-2 clinical isolates. The volume of inoculum was chosen according to titration on A549-hACE2 cells and MDA-MB-231-hACE2 cells aiming to reach ~10% viability of untreated target cells at the time point of harvest. 48 h (MDA-MB-231-hACE2 cells) or 72 h (A549-hACE2 cells) after infection, analysis of virus-induced killing was performed by measurement of viability of target cells using the CellTiter-Glo 2.0 reagent (Promega). Cells were treated according to the manufacturer’s instructions. In brief, 10 µl CellTiter-Glo 2.0 reagent was added to each well, incubated for 10 min in the dark at room temperature and luminescence was recorded using the Infinite F200 microplate reader (Tecan). Viability of cells was calculated by normalization of readings for infected cells relative to those for untreated control cells. Curve fitting and EC50 calculation was done using GraphPad Prism (non-linear regression).
